# Optimizing the antiretroviral treatment focusing on long-term effectiveness and a person-centred approach. Consensus Guidance Using a Delphi Process

**DOI:** 10.1101/2022.05.25.22275575

**Authors:** Franco Maggiolo, Sergio Lo Caputo, Stefano Bonora, Marco Borderi, Antonella Cingolani, Gabriella D’Ettorre, Antonio Di Biagio, Simona Di Giambenedetto, Cristina Gervasoni, Giovanni Guaraldi, Paolo Maggi, Lucia Taramasso

## Abstract

**Background:** Modern antiretroviral therapy (ART) offers several treatment options characterized by high efficacy and tolerability, and new strategies with new drugs are now available for the treatment of HIV infection. As definitive data on the long-term success of these new strategies are lacking, a panel of infectious diseases specialists was convened to develop a consensus on how to tailor and follow in time a person-centered ART approach.

**Methods:** Panelists used a Delphi technique to develop a list of statements describing preferred management approaches for ART and patient monitoring and quality of life evaluation. Panelists provided level of agreement and feedback on consensus statements generated and refined them from the first round through 2 subsequent rounds of voting.

**Results:** Ninety infectious diseases specialists from different Infectious Diseases Centres in Italy participated in the consensus process. A consensus was reached on virological and immunological parameters to use to monitor long term efficacy of antiretroviral treatment, while there was no consensus on the use of specific inflammation and immune-activation markers in clinical routine. The panel agreed on the need of an antiretroviral treatment with the lowest impact on bone, kidney and cardiovascular toxicity and on the utility of quality of life monitoring during the standard follow up of people living with HIV.

**Conclusions:** The consensus statements developed by a panel of infectious diseases specialists may provide guidance to practitioners for a person-centered approach aimed at obtaining long-term virological and clinical success for people living with HIV.

**Key points:** A panel of experts in the care of HIV infection expressed their consensus on which could be the best strategy to achieve and maintain long-term effectiveness in course of antiretroviral therapy, using Delphi methodology.

## Background

Modern antiretroviral therapy (ART) is characterized by several treatment options with high efficacy and tolerability, available for both ART-naïve and -experienced people living with HIV (PLWH)[1,2]. Moreover, new ART strategies with new drugs have been recently introduced in clinical practice and are becoming widely used worldwide [3–8], outlining a scenario where therapies can be increasingly tailored to combine efficacy and tolerability with long-term durability [1,2,9]. The new drug class of integrase inhibitors (INSTIs) has allowed to expand strategies for initiating and simplifying antiretroviral therapy (ART) [3,4,8], but literature data, mainly derived from the clinical practice, highlight how INSTIs cannot yet be considered therapies for every patient [10–14]. Indeed, clinical and virological characteristics of each single subject should be considered before choosing drugs either for ART-naïve or -experienced PLWH. In particular, late presentation, tolerability to previous drugs, patient’s lifestyle, drug-drug interactions, viral subtype, history of previous virological failures or transmitted resistance and even residual viremia can play a role in the maintenance of the long-term efficacy of ART [15,16]. In addition, definitive data on the long-term success of the newest therapeutic strategies are still lacking due to their recent introduction in clinical practice, and the best way to study them throughout time, is still object of debate and research. For instance, it is still to define the role of very low-level HIV viremia, the utility of studying viral reservoirs or markers of immune-activation or subclinical inflammation other than CD4-T and CD8-T cell count in clinical practice for long-term monitoring of ART, and which differences, if any, exist among different ART strategies.

At the same time, the question remains on which treatment could be more adequate in challenging scenarios such as the advanced naïve or AIDS presenter patients, as well as the non-adherent or heavily treatment experienced patients.

New drugs, with lower rates of adverse events and better long-term tolerability compared to past, together with their lower potential for drug-drug interactions or altered metabolism [17] may allow easier management also in the most challenging scenarios, and better ease of intake for PLWH, who consequently may experience an additional benefit in terms of quality of life. Also, the opportunity for single tablet regimens seems to satisfy patient preferences and improve adherence [18–24].

Indeed, considering the patient’s preferences is another key point for the development of tailored person-centered ART strategies, and for the success of long-term therapy. Patient reported outcomes measures (PROMs) allow assessing important aspects of health, beyond immunovirological results, by accessing people’s subjective perceptions of their health-related experiences, which may differ from those perceived by service providers and physicians [25]. Thus, their routine use might be considered to optimize the follow up of PLWH, but also the resource allocation, by focusing on the true perceived needs of people who access services and therapies.

As conclusive data are still not available on which could be the best treatment monitoring strategy during ART, we identified four thematic areas to be submitted to a panel of experts in the care of HIV infection. We used the Delphi methodology to reach a consensus on how to monitor I) clinical efficacy of ART, II) immunology and pharmacology issues, III) long-term safety of ART and IV) how to use quality of life measures and patients reported outcomes in the clinical context.

## Methods

### Study Design

We employed a Delphi technique to develop a list of statements describing optimal management approaches antiretroviral therapy prescription and therapeutic success monitoring. The Delphi technique is a structured process to develop consensus on a topic with gaps in data-driven guidance [26]. Delphi poll was submitted to a restricted panel of expert clinicians working primary as infectious diseases specialists in the care of HIV infection and its complications in different infectious disease centers in Italy.

### Study participants

Panelists were identified based on a combination of clinical experience with managing HIV infection, relevant peer-reviewed publications, and active engagement in professional society meetings. The research team discussed the possibility of joint participation of doctors and PLWH in the implementation of the consensus, but after a discussion on the specificity of some of the issues addressed, the risk of methodological bias by including the assessment of PLWH for only some (and not all) of the elements, and the need for uniformity of expertise to validate the Delphi methodology, the research team decided to carry out the voting only by including doctors specialised in infectious diseases and in the treatment of HIV infection. The reason why other specialists and general practitioners were not included in the panel is that, in Italy, starting in 1990, Law 135/90 [27] introduced a plan of care objectives, implementation of interventions and training to deal with the AIDS emergency, which shifted HIV care from the community to outpatient clinics run by infectious disease specialists. As a result, currently, the management of chronic HIV infection is almost totally hospital-centered with limited involvement of health care professionals other than infectious diseases specialists.

Participation in the study was proposed to 111 physicians from 17 Italian regions; 90 specialists agreed to participate and expressed their level of consent to the submitted statements. Expert panelists were surveyed for their clinical opinions based on their previous experiences and interpretation of the available literature by deploying several rounds of questionnaires. The statements proposed in the questionnaire were formulated by a research team to address clinical, laboratory and therapeutic issues for which data from the literature are still scarce. As the aim of the survey was to find a consensus targeted on modern ART strategies, the focus was on INSTI-based two-drug regimens (2DR) and tenofovir alafenamide (TAF)-based triple therapies, leaving out of this consensus the issues related to other and older dual therapies or 3-drug ART combinations with other backbones. The research team did not participate in the Delphi process and was aware of the identity of the participants but was blinded to the identity of their responses.

### Delphi Round 1

In round 1, levels of aggregation of the consensus were assessed on 20 items selected by the re-search group with the aim of managing the long-term effectiveness of ART with a person-centered approach. Statement 1-5 were focused on treatment monitoring during ART, statement 6-10 on immunology and pharmacology issues, statement 11-14 on long-term safety of ART and statement 15-20 on quality of life and outcomes reported by PLWH. The panelists were asked to express whether they disagreed, partially agreed, agreed or strongly agreed with the proposed statements. For each item, the consensus was considered to have been reached if at least 75% of the voters stated that they agreed or strongly agreed with the statement. The same threshold of 75% was also required for disagreement (disagree or partial agree) to define the consensus against one given statement. All panelists had the possibility of leaving feedback for each statement, that was reviewed blindly by the research team of the study after each round of voting. For all the statements which did not reach the consent, the research team reformulated the statement after revision of the literature on the topic and of the feedback given by the panelists after the first round of voting.

### Delphi Round 2

In round 2, the research team resubmitted the statements which did not reach a sufficient consent in round 1 to the panelists, after rephrasing and content revision. Also in round 2, at least 75% of panelists who declared they agree or strongly agree to the statements, was required to define the consensus was reached. The panelists had the possibility of leaving feedback for each statement. Statements who did not reach the consent were modified again by the investigators based on the feedback.

### Delphi Round 3

In the third and final round, the statements that did not reach consent in rounds 1 and 2 were presented again to the panelists after further revision by the research team, based on the feedback given by the panelists in the previous rounds. At the conclusion of the third round, the investigators considered the consensus was reached in case ≥ 75% of panelists agreed (strongly agree, agree) to the statement. Otherwise, the research team declared that a consensus was not reached for the given item.

## Results

### Characteristics of Panel Members

One hundred and eleven potential panelists were contacted, of which 90 agreed to participate and expressed their grade of consent to the statements identified by the research team.

Among 90 panelists, eight choose to remain anonymous during voting and revision of the statements; among the remaining 82, 27 (33%) were female. Fourteen non-anonymous panelists (17%) were under 40 years old, 34 (41.5%) were between 40 and 55 and 34 (41.5%) were older than 55 years old. All were infectious diseases physicians specialized in HIV care. All 90 panelists participated to the first round of voting, 76 to the second round, and 64 to the third round.

During the three rounds, panelists developed a consensus on treatment monitoring strategy during ART, how to manage immunology and pharmacology issues, long-term safety of ART and use of quality of life measures in the clinical context.

### Clinical efficacy: ART-monitoring strategy

The panelist agreed at first round voting that the aim of ART is to achieve virological success, obtaining not only an HIV-RNA <50 copies/mL during treatment but also, hopefully, an undetectable value of HIV viral load (target not detected, TND). This goal is, at present, achievable and can be maintained in all patients independently from baseline HIV-RNA with the use of triple ART with recognized efficacy and high genetic barrier. The panelists also agreed that an HIV-RNA of at least <200 copies/mL is crucial to bring virus transmission to zero.

Modern therapies, due to their better tolerability and improved drug-drug interaction profile, make the need for therapeutic changes less frequent than in the past. A consensus about clinical monitoring of viral reservoirs such as HIV-DNA and residual viremia was reached after two rounds of voting. In the final statement, it was clarified that the study of viral reservoirs should not be done routinely, but can, in particular clinical situations, be useful in patient’s management [28–31] (Supplementary materials). The panel did not reach an agreement about the utility of using inflammation or immune-activation markers in daily clinical practice. In fact, the study of such markers has been proposed by some panelists to further refine decision-making algorithms, with the aim of defining safer and more tailored therapies [32–38]. However, the clinical relevance of these markers is currently unproven, leading most respondents to express disagreement with their use in clinical practice. On the third and final round of voting, the majority (67%) of voters expressed disagreement (disagree or partial agree). Then, a 75% consent was not reached about this specific item, suggesting that further studies will be needed to clarify the issue (Supplementary materials).

The final statements are reported in Figure 1.

**Figure 1.**
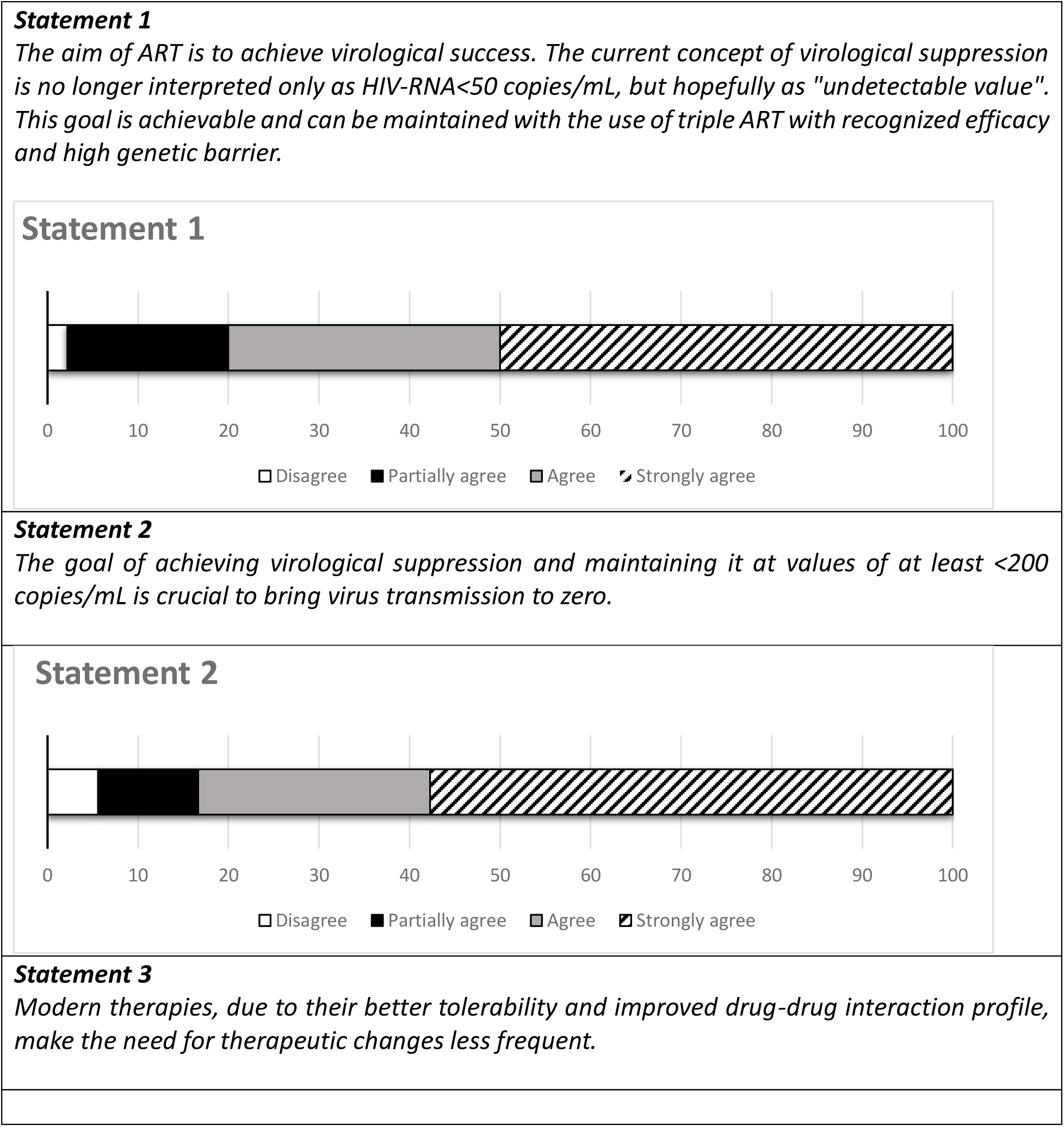
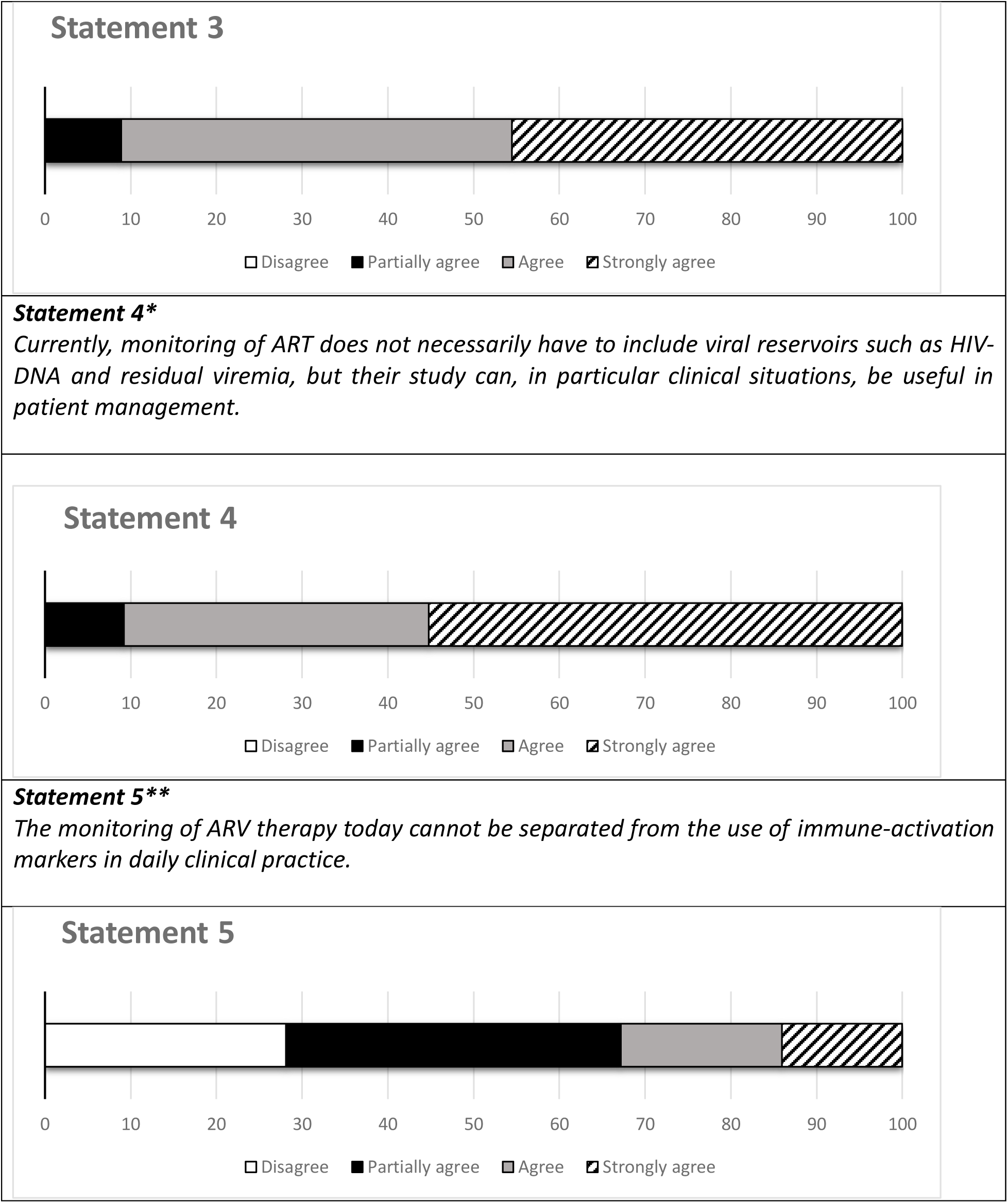
Statements on monitoring long-term therapeutic efficacy. * Agreement was reached for this statement after reformulation and two rounds of voting (Supplementary Figure 1). ** An agreement of at least 75% of the voters has not been reached for this statement, after reformulation and three rounds of voting (Supplementary Figure 2).

### Immunology and Pharmacology

The CD4+ T-lymphocyte (CD4) count remains the most validated immunological prognostic marker according to almost all the study participants, being considered the strongest predictor of clinical progression to AIDS and non-AIDS-related events. Then, the panelists agreed on the utility of its monitoring along time, in conjunction with the assessment of CD4 percentage, which can be less influenced by total white blood cell variability than the absolute CD4 count, and with CD4/CD8 ratio, which can be interpreted as a useful marker of immune-senescence, immune-activation and inflammation, that complements the overall risk assessment of even non-AIDS-defining events [11, 39-41].

Regarding the discussion of pharmacology issues, the panel questioned if in some clinical situations it might be more appropriate to maintain a regimen with a backbone of two nucleoside reverse transcriptase inhibitors (NRTI) than choosing a 2DR. To reach a consensus on this item, the panel requested a revision of the statement after discussion on which are the most challenging clinical scenarios in which the statement can be true, identifying the setting of low adherence and known archived resistances, or lack of complete clinical and virological history in a PLWH needing an antiretroviral switch, or first-line ART in naïve patients with low CD4 or high HIV-RNA load as the possible situations in which to prefer a 2NRTI backbone combined with a third drug, over other strategic combinations [5–7,18,19,42]. Moreover, in patients with suboptimal adherence or who have discontinued therapy one or more times, exploiting the forgiveness of triple therapy may be advantageous in limiting the risk of virological failure and drug resistance mutations [43]. Therapeutic drug monitoring (TDM) could be used as an additional tool to optimize the antiretroviral treatment in complex or special patient populations [44-47].

The final statements are reported in Figure 2.

**Figure 2.**
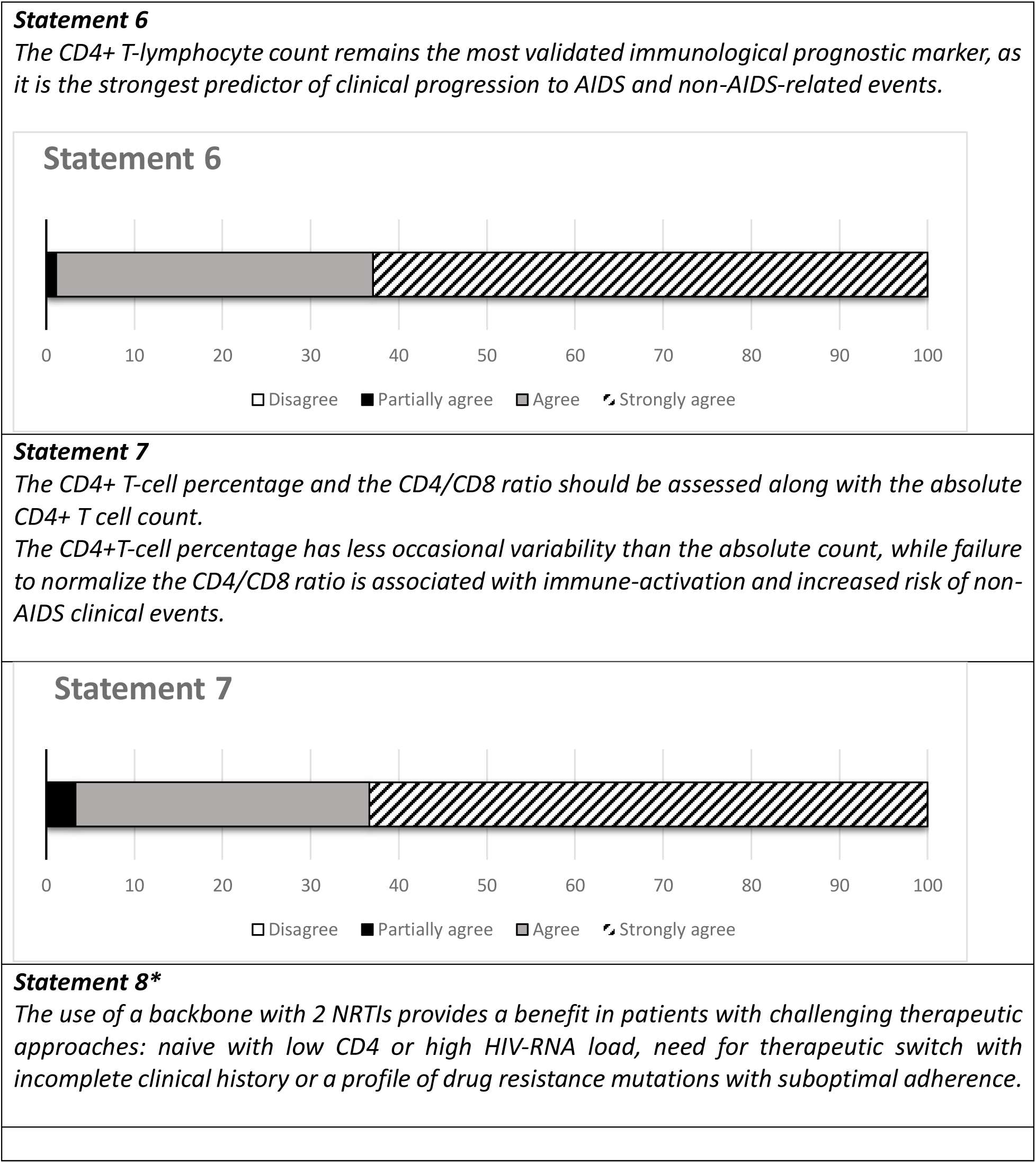
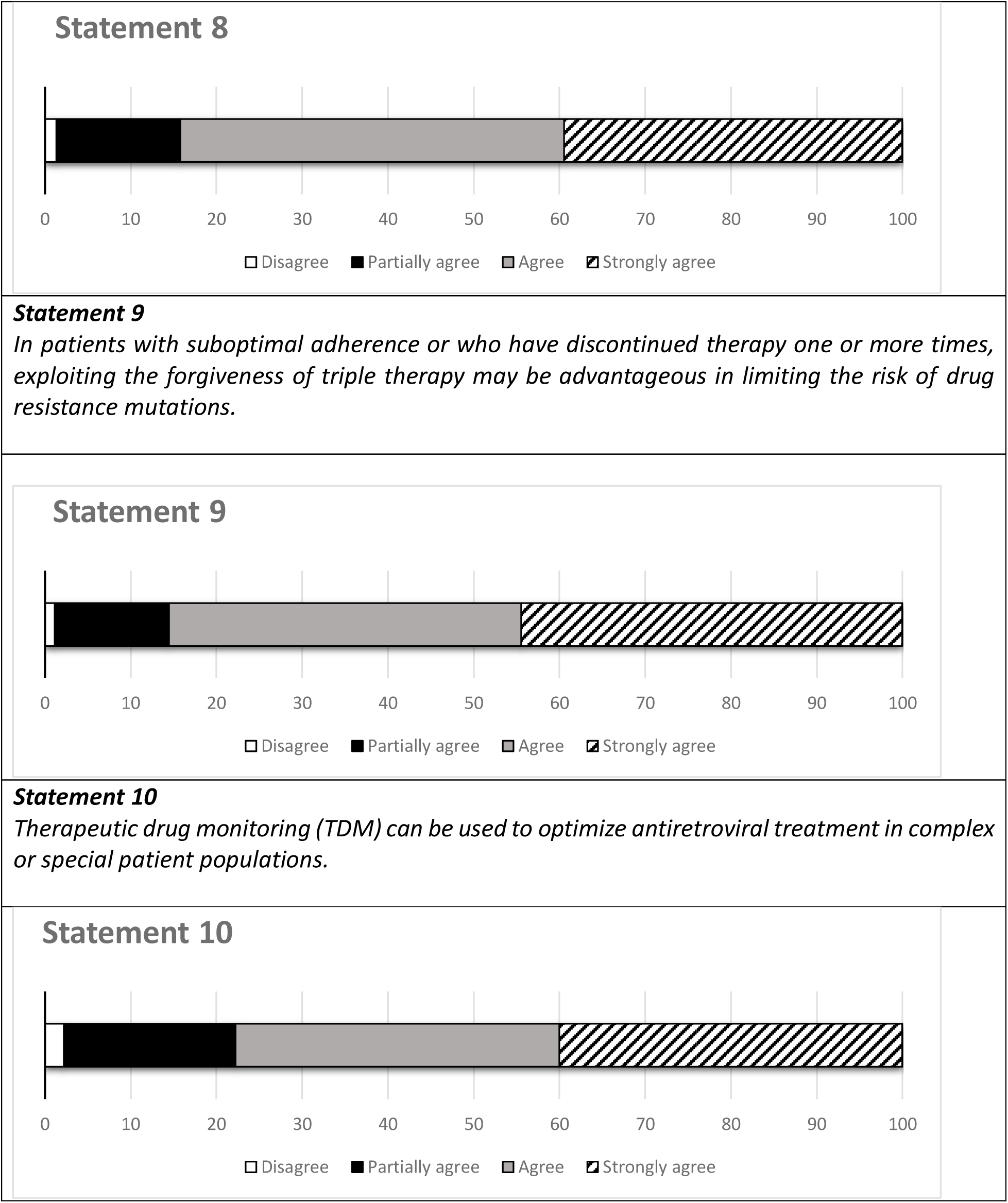
Statements on immunology and pharmacology. * Agreement was reached for this statement after reformulation and two rounds of voting (Supplementary Figure 3).

### Long-term safety

Preventing long-term side effects of antiretrovirals remains a key issue for the success and durability of an ART regimen, also in the light of ageing of PLWH and consequent possible increase of comorbidities. The panel identified three main domains in the field of ART related toxicities to manage: bone, kidney and cardiovascular disease (CVD). These toxicities were mainly linked to the use of older antiretrovirals and thus the possible impact of new therapies was discussed. The discussion also addressed the differences between TAF and tenofovir disoproxil fumarate (TDF) in terms of tolerability and safety.

A consensus was reached among voters, that in the physiological, steady, loss of bone mineral density (BMD) that happens with ageing, a TAF-based triple ART showed no BMD reduction after the first year of therapy. TAF-based triple therapy should also be considered kidney friendly, while special attention and monitoring should be reserved for patients with previous or current renal damage or concurrent use of nephrotoxic drugs.

Regarding CVD, the panel discussed two items: how to evaluate the risk in PLWH who experience change in lipid levels during ART and how to deal with the weight gain that has been observed during treatment with new drugs such as INSTIs and TAF. The consensus was reached about the indication to assess CVD risk in PLWH on TAF-based ART using up-to-date risk scores (e.g. Framingham, ASCVD, D:A:D:) and not only on the basis of change in plasma lipids. On the other side, the panelists required three rounds of voting and statement revision to specify that the weight gain observed with modern drug regimens did not show correlation with increased CVD risk in people with a body mass index (BMI) <30 kg/m^2^ and in absence of metabolic syndrome and other risk factors, but they also required to include in the statement that a longer follow-up is needed in order to provide more precise indications, also considering the multiple concomitant factors that might influence weight gain in course of ART (Supplementary materials). The final statements are reported in Figure 3.

**Figure 3.**
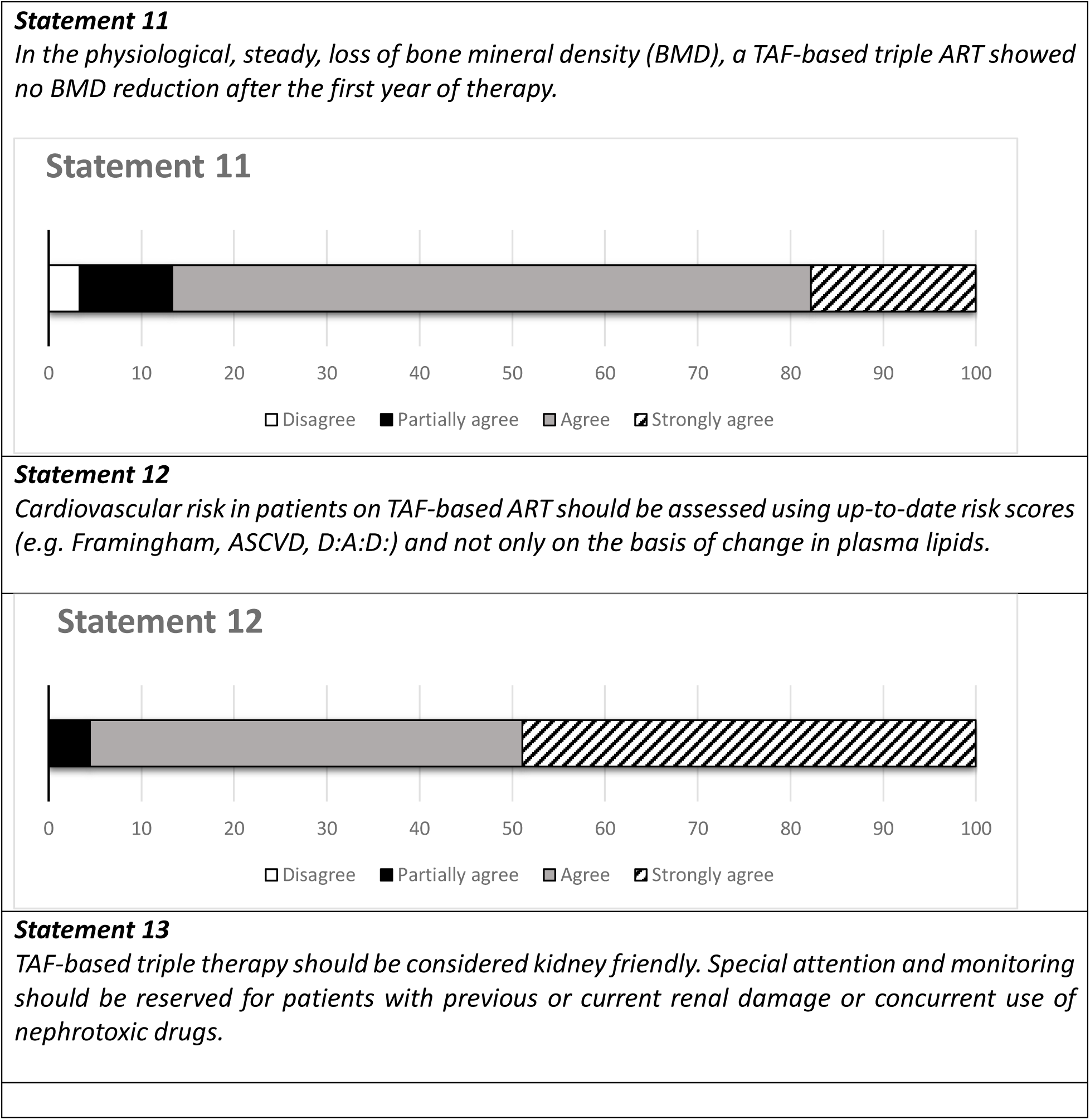
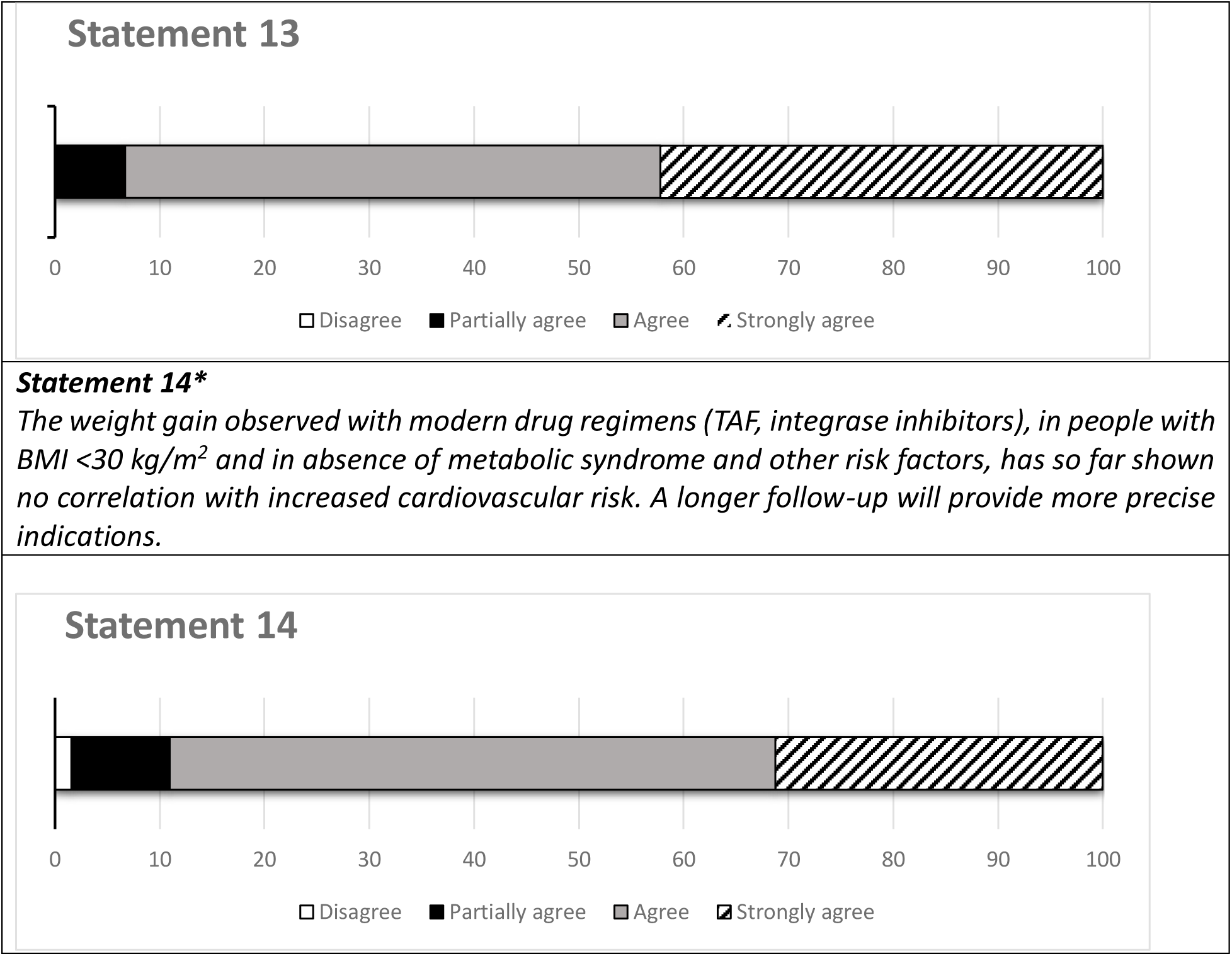
Statements on long-term safety of antiretroviral therapy (ART). * Agreement was reached for this statement after reformulation and three rounds of voting (Supplementary Figure 4).

### Quality of life

According to the panelist judgements, health related Quality of Life (HRQoL) should be assessed routinely in the clinical management of PLWH. Improving the HRQoL of PLWH has the potential to improve several clinical outcomes, including medication adherence, retention in care and well-being. HRQoL should be measured with PROMs validated in PLWH, with the aim of achieving a person-centered approach, but also of using resources efficiently, facilitating discussion of sensitive issues with PLWH, encouraging a greater engagement with services, and promoting adherence to treatment.

The study participants agreed that HRQoL in PLWH does not depend only on ART, but also on multifactorial causes both related to health status (such as multimorbidity, frailty, symptom burden, disability) and to the social context (such as social isolation, financial insecurity, HIV-related stigma and discrimination), indicating the need for people-based health care models. There are no conclusive data demonstrating a significant impact of different ART regimens on HRQoL, but individual domains describing HRQoL may be differently affected by specific antiretroviral drug-related toxicities. Moreover, the consensus was reached that HRQoL reported by PLWH is better in single tablet regimens (STR) than in multi-tablet regimens and that HRQoL monitoring over time assesses the experience of drug burden (number of tablets, food restrictions, ease of intake) and symptoms associated with treatment regimes. The complexity of the pharmacological burden, identifiable in the definition of polypharmacy, entails consequences for care, but also ethical, social and economic, while the concepts of deprescribing and therapeutic optimization might allow the reduction of potentially inappropriate drugs and drug-drug interactions, with the ultimate aim of improving the patient’s quality of life [48-51].

The final statements are reported in Figure 4.

**Figure 4.**
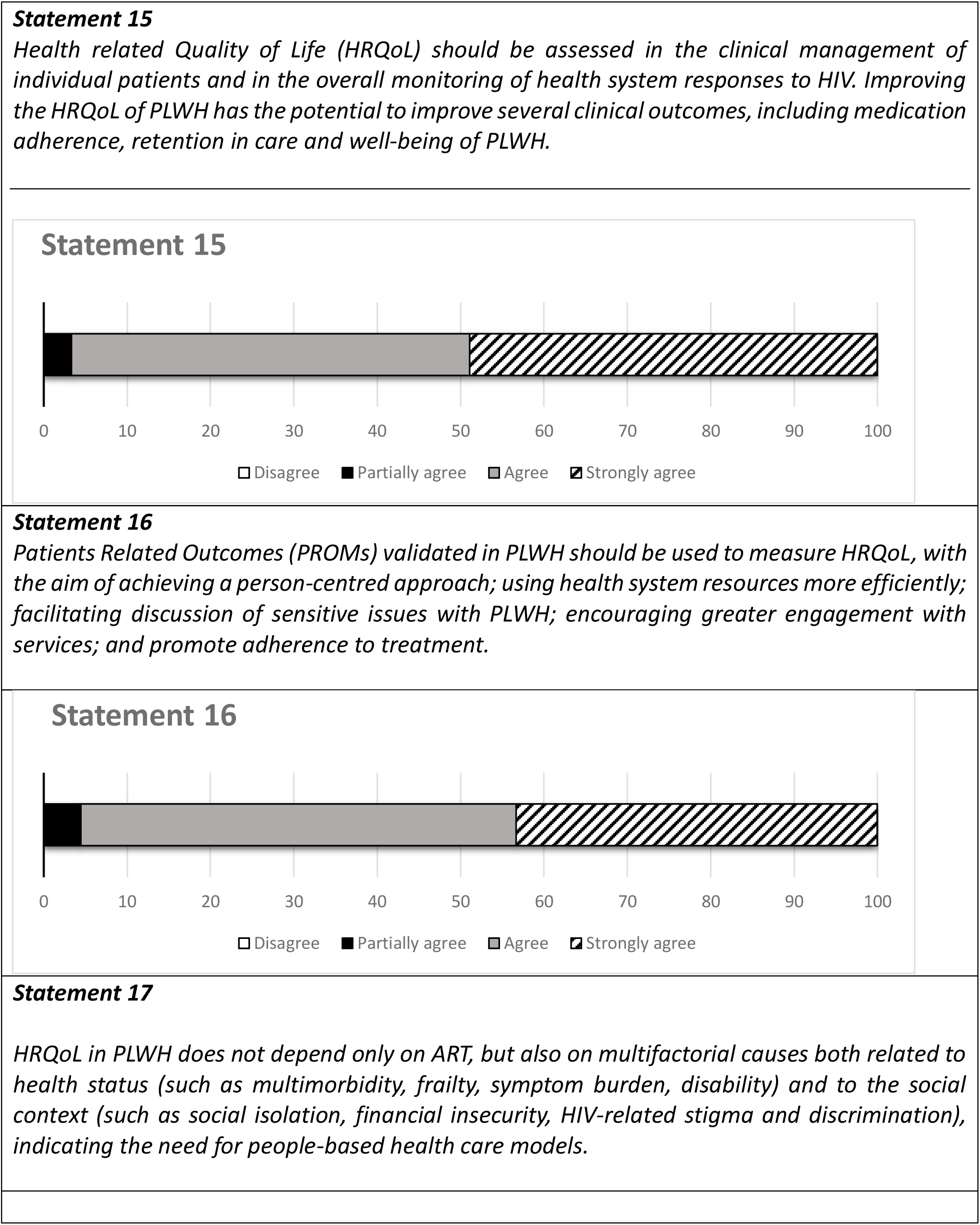
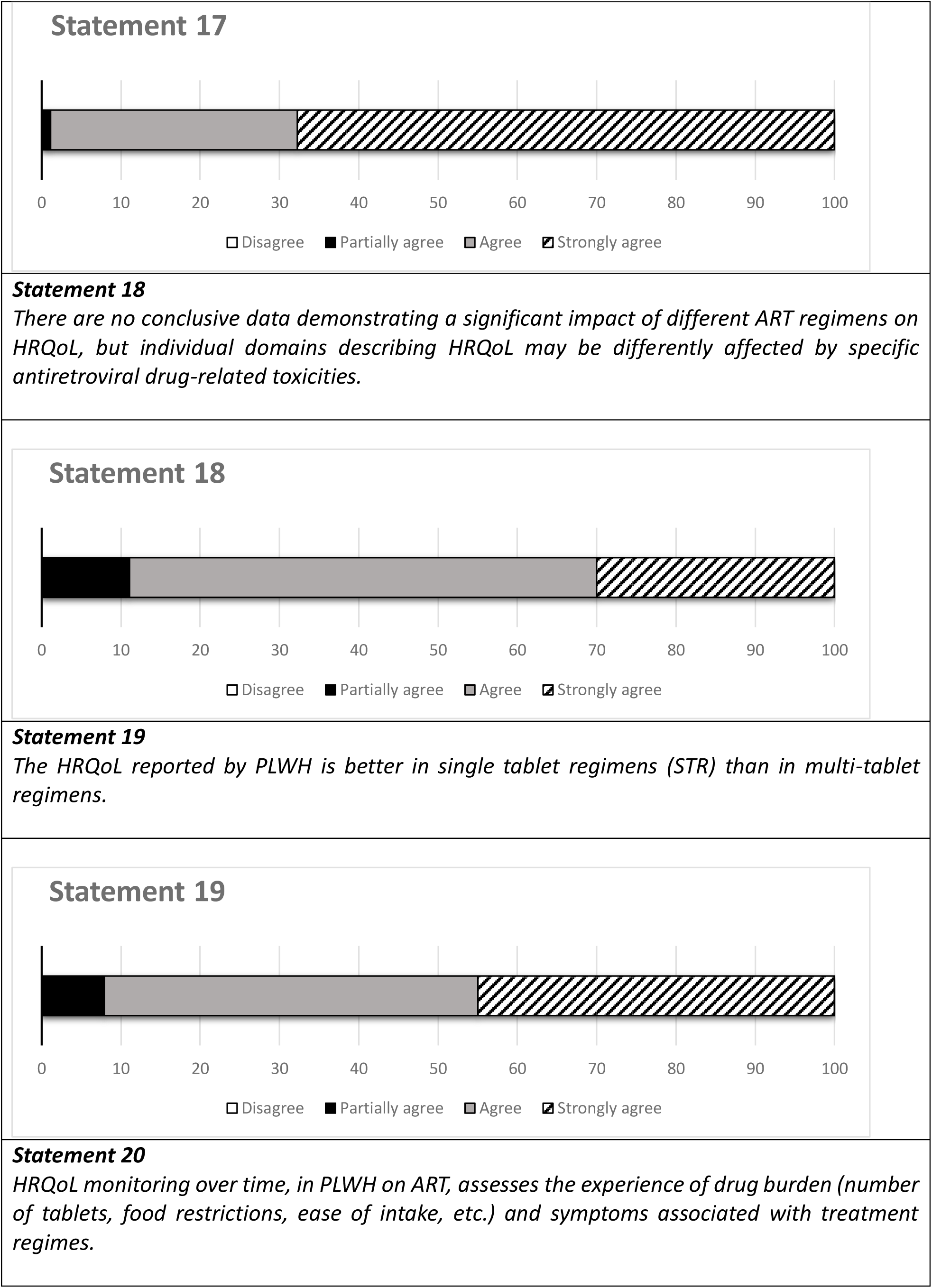
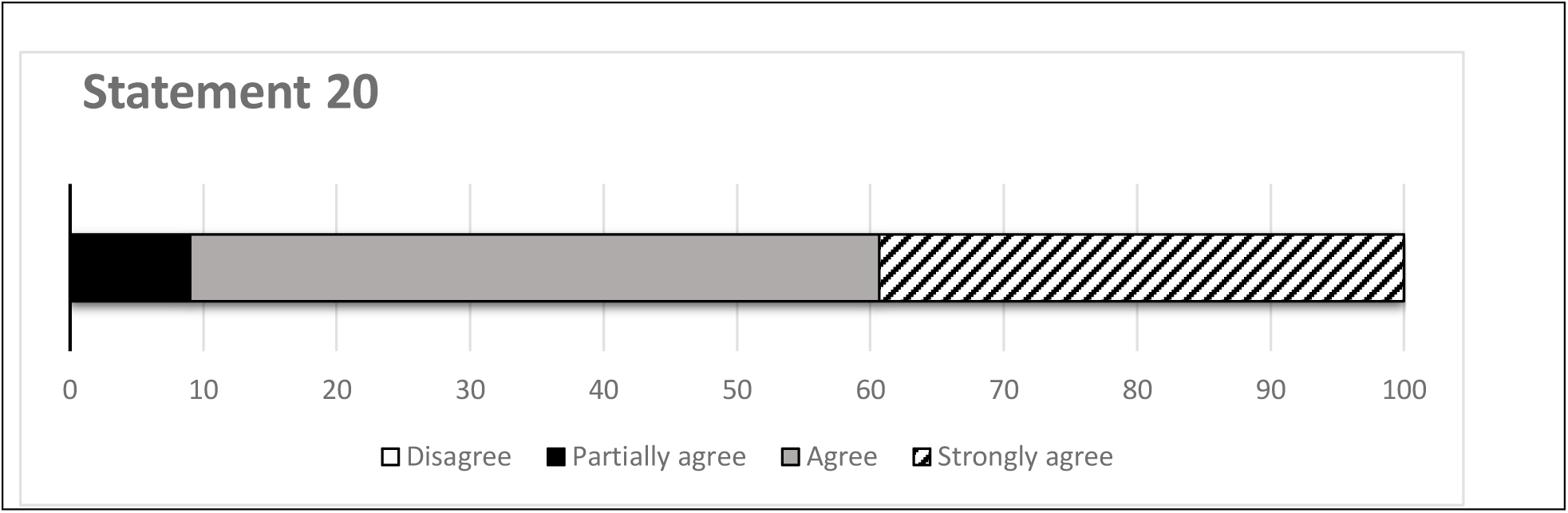
Statements on quality of life assessment.

## Discussion

In this study, a panel of 90 experts in the care of HIV infection expressed their consensus on which could be the best strategy to achieve and maintain long-term effectiveness of antiretroviral therapy. The agreement was reached on virological and immunological parameters to be used to monitor antiretroviral therapy, pointing out how, beyond the assessment of HIV-RNA, the monitoring of CD4 count and percentage, remains crucial to assess the risk of AIDS and non-AIDS related events [52-57]. The evaluation of CD4/CD8 ratio was felt remarkably relevant as well, as a marker of immune-senescence, immune-activation and inflammation [11, 39-41], while the direct study of pro-inflammatory cytokines or other markers of immune-activation and inflammation was generally considered by the panelists an object of research rather than a tool in clinical practice.

The panelists agreed that modern therapies allow to bring together high genetic barrier, low potential for drug-drug interactions or altered pharmacokinetics, and good tolerability, with different drug combinations being available in single tablet regimens (STRs), too [5–7,18,19,42]. In this context, the forgiveness of an ART, namely its the ability of achieving and sustaining viral suppression, despite suboptimal adherence [58-60], was considered as an additional feature which can further improve possibility of long-term success. Few data are available comparing two-drug (2DR) and triple-drug regimens, however, experimental models in vitro showed higher forgiveness of TAF-based triple therapy compared to 2DR, and consistent protection against emergence of drug resistance during simulations of short lapses in adherence [43], supporting the choice of this strategy in patients with suboptimal compliance. In such situations, the panelists also considered the contribution of TDM to be important. In PLWH, TDM can be used to monitor adherence, but, importantly, also to optimize drug exposure in cases in which the knowledge of the pharmacokinetics of antiretroviral drugs is incomplete, such as in cases of insufficient excretory organs, dialysis procedures, or possible altered absorption or distribution of the ART, such as in gastro-resected PLWH, those undergoing polypharmacy or obese patients or pregnant women [44-47]. Then, the panel concluded that TDM might be key to optimize antiretroviral treatment in complex or special patient populations, although unfortunately, until today, it is not widely accessible in routine clinical practice in many infectious diseases centres.

Another point considered key for the long-term success of ART is the prevention of long-term side effects [61], in consideration also of ageing and co-morbidities of PLWH [62,63].

The panelist agreed that, in recent years, the introduction of new drug classes and regimens has radically improved the long-term tolerability, having no effects on plasma lipid increases, and no interference with renal, bone and cardiovascular outcomes, as compared to the older antiretrovirals [1,64–67]. These characteristics have even greater relevance considering that PLWH have already an increased risk of bone [68–70], renal [71], or cardiovascular problems with ageing [72,73], regardless of therapy. Finally, in the most recent years, the new issue of weight gain has emerged. Indeed, studies mostly conducted in Africa, have shown that PLWH who began antiretroviral therapy, especially if ART-naïve, in more advanced stage of HIV disease, with lower baseline body mass index (BMI), and treated with TAF or INSTIs, experienced a significant increase in body weight [74–78]. It remains to be established to what extent this weight gain is attributable to a ‘return to health’ phenomenon and how much to an underlying pathological mechanism, and whether weight gain may increase the CVD risk of PLWH [79]. However, both TAF and INSTIs are lipid-neutral drugs and the panelists therefore considered them suitable treatment options in PLWH at CVD risk [1,2,80–82]. On the other side, a possible and independent increase in CVD risk in parallel with weight gain during these same treatments has no supporting data, nor have switching strategies to other (and potentially less tolerated) regimens, which are not, at present, supported by guidelines or other research data. Longer follow-up and the results of ongoing studies will provide more precise guidance on this issue. The last factor that was discussed in the Delphi consensus was the evaluation of quality of life in PLWH. In 2015, UNAIDS promoted the 95-95-95 agenda to end AIDS as a public health threat by 2030. The ‘fourth 95’ goal was added as the ultimate objective of HIV care: to ensure that 95% of PLWH with suppressed viral load achieve a good health-related quality of life (HRQoL). The relevance of HRQoL as a measure of well-being among PLWH is a well-established concept [83] and can be assessed by PROMs. PLWH report lower HRQoL than the general population and compared to people with other chronic health conditions [84–86]. Lower HRQoL scores were found to be closely related to worse clinical outcomes such as low CD4 counts, hospitalization and the occurrence of AIDS-defining events, [87–91] while high HRQoL scores correlate with positive outcomes, such as treatment adherence and retention in care [92]. While comorbidity, disability and pain compromise HRQoL, successful HIV treatment has a positive impact on it [93]. The panel recognized the evaluation of PROMs as an important tool in the care of PLWH and considered it key in therapeutic strategy studies, where they feel that HRQoL domain should contribute the global evaluation of the effectiveness of a treatment [3,94,95]. HRQoL monitoring into clinical and community services can provide a direct feedback on the person-centred perspective on the effectiveness of interventions, has the potential to identify PLWH who would benefit from specific services and to address modifiable factors that negatively impact on HRQoL [96].

The results of this Delphi consensus have some limitations to consider. Due to the nature of the Delphi process, the panel addressed issues for which data from the literature are very scarce, and therefore it should be noted that the final statements that were presented reflect the opinion of a group of experts in the field, rather than evidence-based guidance. Furthermore, the panelists were limited to infectious disease specialists involved in HIV care, but as also emerged in this consensus, modern ART management cannot be done without including patients’ opinions in the decision-making process. Consequently, due to the many technical aspects covered in the statements, it was decided to carry on the work only among infectious disease specialists.

Even with the limitations mentioned above, this consensus addressed many challenges of modern ART, concluding that the newer regimens do offer significant advantages over older treatment options and that new monitoring strategies including TDM and PROMs evaluations may bring closer to the goal of personalised therapies. The consensus statements developed in this study can provide guidance to practitioners in building a person-centred approach aimed at achieving long-term virological and clinical success for PLWH.

## Data Availability

All data produced in the present work are contained in the manuscript

## Data Availability Statement

All data on the rate of agreement with the statements proposed in the Delphi consensus are provided in the text and supplementary materials.

## Patient Consent Statement

Not applicable.

## Acknowledgments

**Delphi reviewer committe:** Massimo Andreoni, Università degli Studi di Roma “Tor Vergata”, Roma; Andrea Antinori, INMI “Lazzaro Spallanzani” I.R.C.C.S., Roma; Giovanni Di Perri, Ospedale Amedeo di Savoia, Università degli Studi di Torino; Andrea Gori, Fondazione IRCCS Ca’ Granda Ospedale Maggiore Policlinico, Milano; Cristina Mussini, Azienda Ospedaliero-Universitaria Policlinico di Modena; Giuliano Rizzardini, ASST Fatebenefratelli Sacco - Ospedale Luigi Sacco, Milano; Antonella d’Arminio Monforte, ASST Santi Paolo e Carlo - Polo Universitario, Milano; Antonella Castagna, IRCCS Ospedale San Raffaele Università Vita-Salute, Milano.

**Study group:** M. Allegrini, G. Angioni, L. Attala, F. Bai, A. Bandera, O. Bargiacchi, A. Beltrame, B. Borchi, M. Brizzi, A. Calcagno, M. Camici, M. Campus, D. Canetti, S. Casari, A. Cascio, B.M. Celesia, A. Celotti, G. Cenderello, L. Chessa, S. Cicalini, R. Cinelli, M. Colafigli, P. Colletti, N. Coppola, A. Costantini, L. Cristiano, G. Cuomo, M. De Gennaro, P. De Leo, F.G. De Rosa, F. Del Puente, G. Di Giuli, E.M. Erne, V. Esposito, M. Fabbiani, D. Farinacci, S.M. Ferrara, E. Foca’, D. Francisci, A. Franco, F.M. Fusco, I. Gentile, A. Giacometti, N. Gianotti, G. Guadagnino, M. Guerra, A. Ialungo, F. Lagi, M. Lanzafame, G. Lapadula, A. Lazzaro, A. Longo, G. Madeddu, E. Manzillo, A. Masiello, M. Menozzi, B. Menzaghi, L. Meroni, P. Milini, M.C. Moioli, A. Muscatello, G. Orofino. F. Ortu, S. Ottou, G. Pantò, A. Parisini, G. Parruti, A. Patacca, T.S. Prestileo, D. Ripamonti, S. Rusconi, R. Scaggiante, P. Scerbo, M.P. Sciotti, M. Scuderi, L. Sighinolfi, L. Taramasso, E. Teti, C. Torti, M. Trezzi, J. Vecchiet, D. Vitullo.

