## Supplementary Figure 1. Supplementary Figure 2. Supplementary Figure 3. for "Optimizing the antiretroviral treatment focusing on long-term effectiveness and a person-centred approach. Consensus Guidance Using a Delphi Process"

Supplementary materials

For those statements for which there was no consensus in the first round of voting, the statement was reworded and resubmitted. Below are the original texts submitted and the subsequent amendments until the consensus of the voters was obtained.

**Figure 1.** Statement 4

***Statement 4- First round***

*Monitoring of ARV therapy today should include viral reservoirs such as HIV-DNA, residual viremia, and so on.*

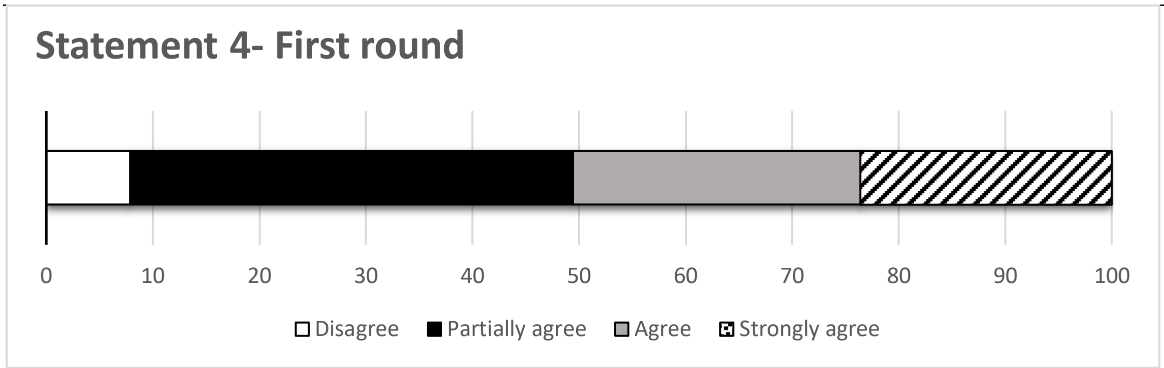


***Statement 4- Second round***

*Currently, monitoring of ART does not necessarily have to include viral reservoirs such as HIV-DNA and residual viremia, but their study can, in particular clinical situations, be useful in patient management.*

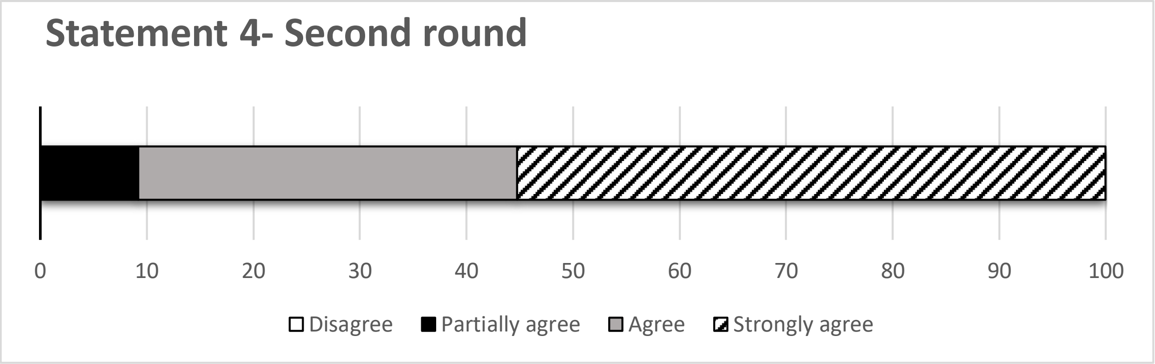

**Figure 2.** Statement 5

***Statement 5- First round***

*Monitoring of ARV therapy today should include markers of immune-activation such as pro-inflammatory cytokines.*

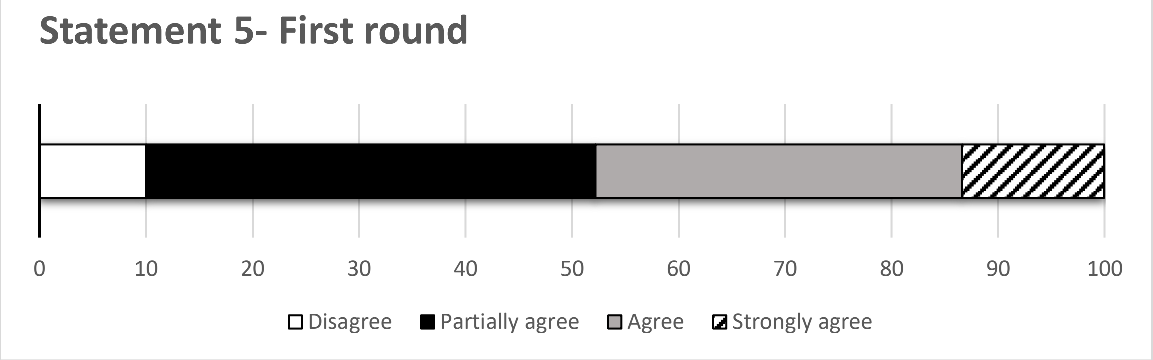


***Statement 5- Second round***

*Monitoring ARV therapy today cannot be done without the use of markers of immune-activation, such as pro-inflammatory cytokines.*

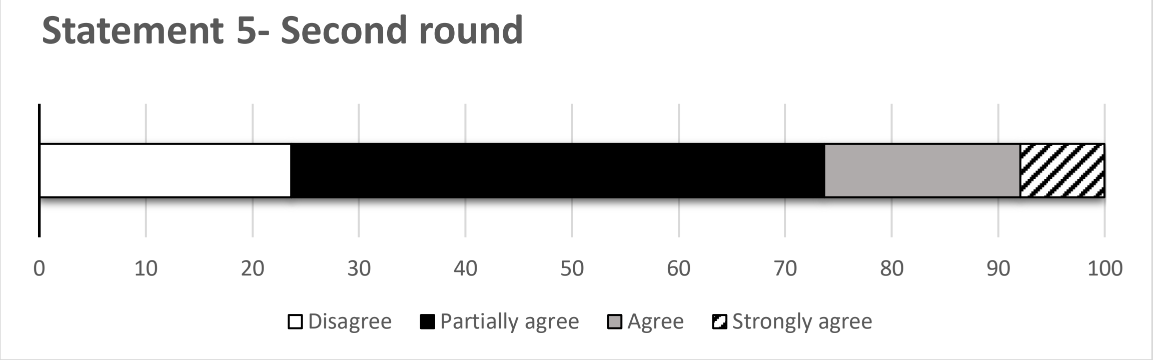


***Statement 5- Third round***

*The monitoring of ARV therapy today cannot be separated from the use of immune-activation markers in daily clinical practice.*

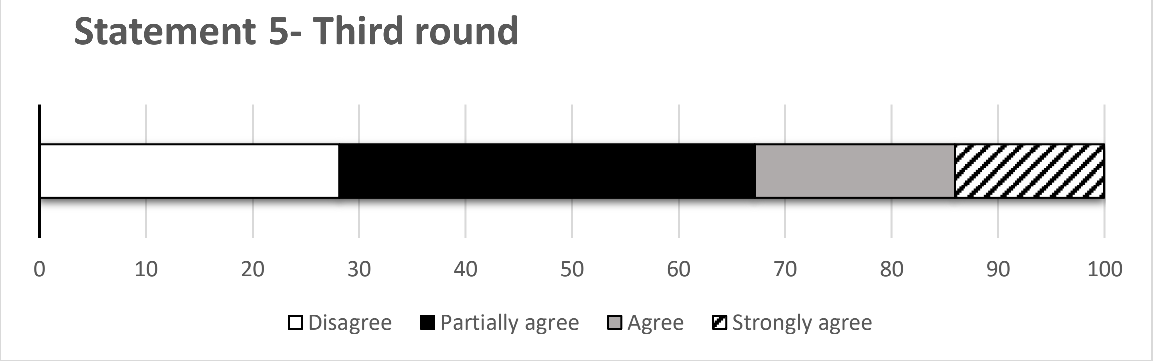

**Figure 3.** Statement 8

***Statement 8- First round***

*Patients with challenging therapeutic approaches (e.g. low CD4, impaired pharmacokinetics, suboptimal efficacy due to previous resistance) may benefit from a backbone based on tenofovir (either TDF or TAF) and emtricitabine.*

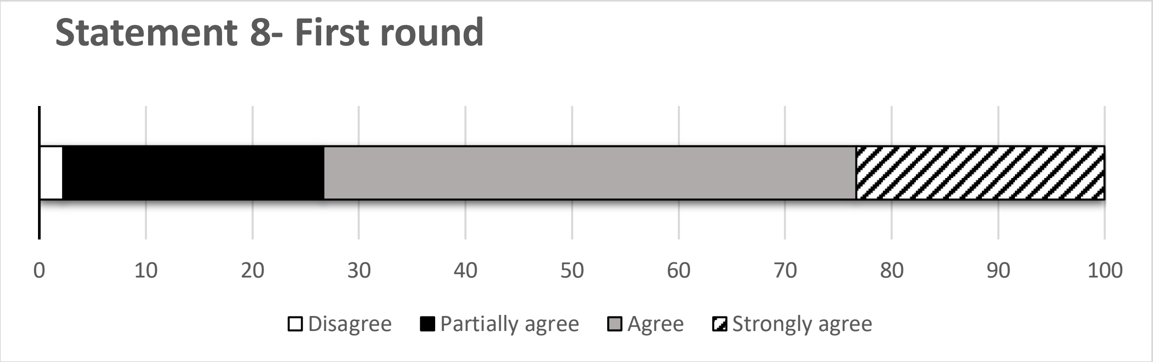


***Statement 8- Second round***

*The use of a backbone with 2 NRTIs provides a benefit in patients with challenging therapeutic approaches: naive with low CD4 or high HIV RNA load, need for therapeutic switch with incomplete clinical history or a profile of drug resistance mutations with suboptimal adherence.*

**Figure 4**. Statement 14

***Statement 14- First round***

*The weight gain observed in TAF-based regimens currently has no correlation with increased cardiovascular risk.*

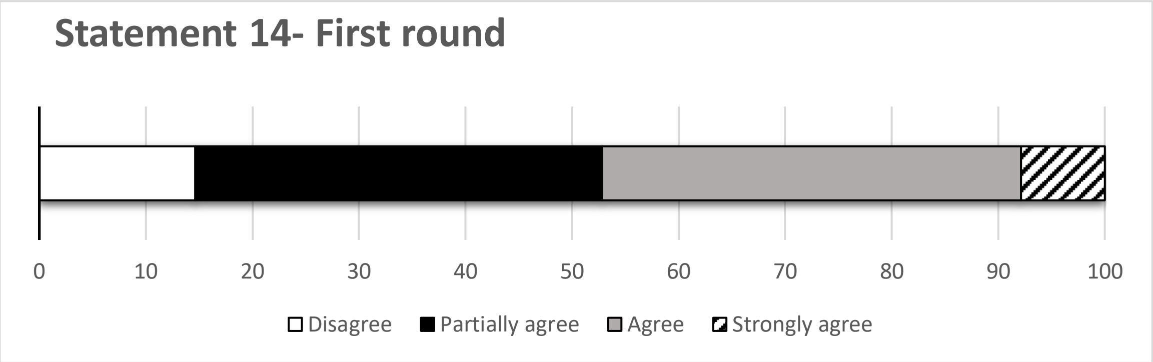


***Statement 14- Second round***

*The weight gain observed with modern drug regimens (TAF, integrase inhibitors), in people with BMI <30 kg/m$^2$ and in absence of metabolic syndrome, has so far shown no correlation with increased cardiovascular risk.*

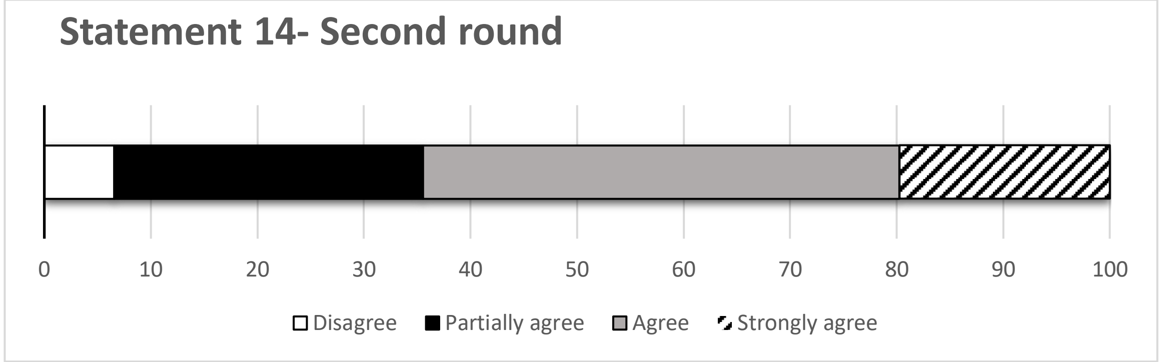


***Statement 14- Third round***

*The weight gain observed with modern drug regimens (TAF, integrase inhibitors), in people with BMI <30 kg/m$^2$ and in absence of metabolic syndrome and other risk factors, has so far shown no correlation with increased cardiovascular risk. A longer follow-up will provide more precise indications.*

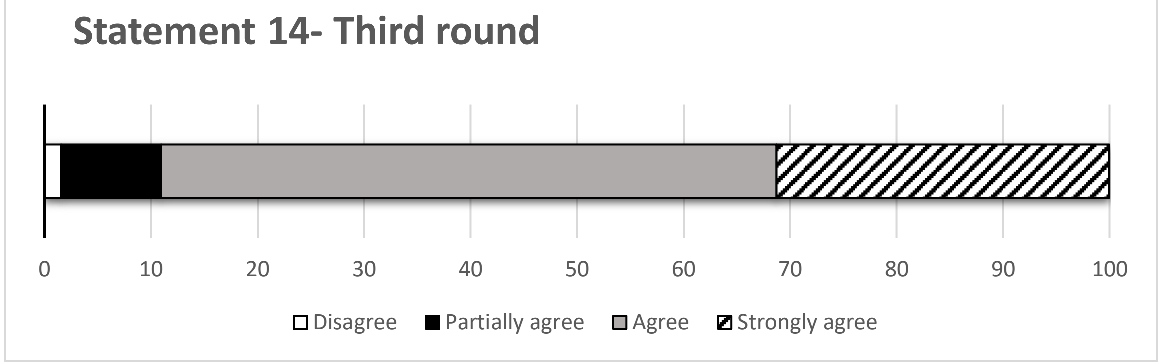
